# Validation of expert system enhanced deep learning algorithm for automated screening for COVID-Pneumonia on chest X-rays

**DOI:** 10.1101/2020.10.20.20213793

**Authors:** Prashant Sadashiv Gidde, Shyam Sunder Prasad, Ajay Pratap Singh, Nitin Bhatheja, Satyartha Prakash, Prateek Singh, Aakash Saboo, Rohit Thakar, Salil Gupta, Sumeet Saurav, M V Raghunandanan, Amritpal Singh, Viren Sardana, Harsh Mahajan, Arjun Kalyanpur, Atanendu Shekhar Mandal, Vidur Mahajan, Anurag Agrawal, Anjali Agrawal, Vasantha Kumar Venugopal, Sanjay Singh, Debasis Dash

## Abstract

The coronavirus disease of 2019 (COVID-19) pandemic exposed a limitation of artificial intelligence (AI) based medical image interpretation systems. Early in the pandemic, when need was greatest, the absence of sufficient training data prevented effective deep learning (DL) solutions. Even now, there is a need for Chest-X-ray (CxR) screening tools in low and middle income countries (LMIC), when RT-PCR is delayed, to exclude COVID-19 pneumonia (Cov-Pneum) requiring transfer to higher care. In absence of local LMIC data and poor portability of CxR DL algorithms, a new approach is needed. Axiomatically, it is faster to repurpose existing data than to generate new datasets. Here, we describe CovBaseAI, an explainable tool which uses an ensemble of three DL models and an expert decision system (EDS) for Cov-Pneum diagnosis, trained entirely on datasets from the pre-COVID-19 period. Portability, performance, and explainability of CovBaseAI was primarily validated on two independent datasets. First, 1401 randomly selected CxR from an Indian quarantine-center to assess effectiveness in excluding radiologic Cov-Pneum that may require higher care. Second, a curated dataset with 434 RT-PCR positive cases of varying levels of severity and 471 historical scans containing normal studies and non-COVID pathologies, to assess performance in advanced medical settings. CovBaseAI had accuracy of 87% with negative predictive value of 98% in the quarantine-center data for Cov-Pneum. However, sensitivity varied from 0.66 to 0.90 depending on whether RT-PCR or radiologist opinion was set as ground truth. This tool with explainability feature has better performance than publicly available algorithms trained on COVID-19 data but needs further improvement.

## 1 Introduction

The Severe Acute Respiratory Syndrome Coronavirus-2 (SARS-CoV-2) pandemic has created global disruption on an unprecedented scale [38]. Tracing, testing, isolating and treating is the universally agreed upon strategy for limiting the spread and minimizing adverse outcomes, but this sequence often faces a bottleneck at the testing stage. Quantitative Reverse Transcription Polymerase Chain Reaction (qRT-PCR), the gold standard, is a specialized test often not available in a timely manner [37]. Further, infected patients need assessment of severity which qRT-PCR does not provide. This led to advocacy of using ubiquitously available resources like Chest X-rays for identifying likely infections, while awaiting qRT-PCR results, and determining severity of coronavirus disease (COVID-19) pneumonia. The concept of causative diagnosis from X-rays might be concerning for uninitiated medical practitioners. This is due to the conventional approach of understanding of pathologies based on casuality as it also guides the treatment principles. Most deep learning algorithms on the other hand learn patterns of association by analysing large volumes of data thereby offering an opportunity to identify associations hitherto unknown or unexplored by medical practitioners. Several studies have demonstrated that deep learning can make causative diagnosis like tuberculosis from X-rays as good as radiologists [27].

COVID-19 pneumonia (CoV-Pneum) has characteristic findings on Chest X-rays (CxR). Bilateral peripheral hazy or consolidative opacities are seen; usually without pleural effusion or mediastinal lymphadenopathy. These findings overlap with presentations of other atypical or viral pneumonias but are different from the typical bacterial pneumonia with large asymmetric consolidation, air bronchograms and pleural effusion. Experienced radiologists use such distinctions to determine whether an abnormal CxR represents a high likelihood of CoV-Pneum. Non-pneumonia forms of COVID-19 cannot be diagnosed on CxR, at least by the human eye. AI algorithms for COVID-19 prediction on CxR have claimed high performance in determining not only CoV-Pneum, but also SARS-CoV-2 infection without any obvious pneumonia on CxR [3], [24], [10]. Most of these algorithms are deep learning (DL)-based [23], [2], [22], [15], [5] trained on a limited amount of COVID-19 data that is available, and without clear explainability. This leads to three concerns. First, robustness or generalizability is typically low for unexplainable AI, especially when DL systems are trained on small datasets. Second, in small datasets when a subset is set aside for validation, apparent algorithm performance is artificially boosted. Last, many datasets do not make clear distinctions between the RT-PCR diagnosis of SARS-CoV-2 infection and the radiological or clinical one of CoV-Pneum. It is *a priori* unlikely that AI tools can detect SARS-CoV-2 infection that has not manifested in the lung, on a CxR. Thus, it seems reasonable to develop explainable AI tools that merge established DL capacity in detecting routine CxR pathology, with logic-based determination of CoV-Pneum likelihood from nature and spatial distribution of the pathology. Whether such an approach would perform as good as pure DL based methods is not known.

Here, we report an AI classifier of CoV-Pneum that is composed of an ensemble model consisting of three DL modules and an expert decision system. The DL modules include pathology classification, lung segmentation, and opacity detection models and are explainable to the extent of an activation map output. The expert decision system is a rule-based classification system that classifies the X-ray into one of three classes, namely COVID-unlikely, indeterminate, and COVID-likely and is fully explainable as well as modifiable as needed. We tested this classifier on two test sets that had not been seen by the algorithm. First, a general population dataset of randomly selected CxR from an Indian quarantine-center. SARS-CoV-2 infection status was not known for this dataset and a single radiologist had read the films. Second, a curated data set for which SARS-CoV-2 infections status was clearly known and four independent radiologists had classified the probability of CoV-Pneum and marked the lesions. Other than concordance with RT-PCR and radiological diagnosis, we also defined concordance between the bounding boxes of lesions identified by the radiologist and the CovBaseAI algorithm. We found that this approach matches the performance of published DL systems, while providing a high degree of explainability. Given the social and medical consequences of being declared “COVID positive”, this appears to be a better approach.

## 2 Related Work

The advent of deep learning techniques and its applications in the tasks involving classification [13], [11] and object detection [31], [29] has already proven its efficacy which led to the escalation in efforts for the development of deep learning based methods for the screening of SARS-CoV-2 utilizing medical imaging. Multiple studies have been proposed for detection of SARS-CoV-2 employing CT scans and chest X-Rays (CXR) [36], [23], [2], [22], [15], [5], [28]. Leveraging CXR imaging for screening of COVID-19 pandemic has several advantages such as portability and accessibility, specifically in areas having inadequacy of resources and areas which are declared as hot spots for viral infection and thus can aid in triaging of affected patients.

As one of the early efforts, [36] proposed a convolutional neural network based approach for screening of COVID-19 patients. They termed it COVID-Net and trained it using open source COVIDx dataset [6]. COVID-Net incorporates features such as architectural diversity, long range connectivity and a light weight design pattern for classification among normal, COVID-19 and pneumonia CxRs. COVID-Net reported an accuracy of 93.3% on COVIDx dataset, a sensitivity of 91% for COVID-19 and a high positive predictive value (PPV) reflecting few false positives. It also incorporates an explainability mechanism by leveraging GSInquire [18]. In another study proposed by Ozturk et al. DarkCovidNet [23] architecture was presented which is inspired by DarkNet [30], a proven deep learning architecture for high speed object detection. DarkCovidNet is trained on a dataset of 1125 images comprising of COVID-19(+), Pneumonia and No-Findings. It resulted in an accuracy of 98.08% and 87.02% for binary and three classes, respectively. Performance of both the architectures [36], [23] discussed above can be significantly improved provided they are trained on larger dataset and can result in increased generalization. However to overcome the issue of data scarcity Afshar et al. [2] proposed COVID-CAPS based upon capsule networks [33] as conventional convolutional neural networks (CNNs) can lose spatial information between image specimens and need substantially larger datasets to train for better performance. COVID-CAPS on the other hand is capable of handling small datasets. COVID-CAPS is trained on dataset having four labels i.e. Normal, Bacterial, Non-COVID Viral and COVID-19 and obtained an accuracy of 95.7% and specificity of 95.8% which is further enhanced after incorporating pre-training and transfer learning based on additional dataset of X-ray images. Many of these models reported cross-validation accurcay and did not reflect upon the need to test developed algorithms on independent blind datasets based on both the PCR tests and the radiological findings. G. Maguolo and L. Nanni [20] have reviewed the recently published literature on CxR based COVID-19 detection and included most of the recently published datasets which had been utilized for deep learning based COVID-19 detection. They demonstrated that models could identify the source correctly even after the lung fields are masked with black box thereby concluding that most models might be biased and learn to predict features that depend more on the source dataset than the relevant medical condition. For tools to be useful in screening and triaging, all three detection, explainability and grading of disease is needed. Additionally, there are several other investigations on COVID-19 detection employing deep learning based methods utilizing CT scans and chest X-ray images [21], [17].

Although a number of tools and models have been developed and published that claim to detect COVID-19 infection from CxRs, there is still a need of a solution for clinical triaging in resource constrained environment. To address this we propose a robust novel framework comprising coalescence of deep learning architectures and rules devised by radiologists for screening of COVID-19 disease. In the following sections, the proposed methodology is described in detail followed by datasets utilized for training evaluation and the results obtained.

## 3 Methodology

The architecture of CovBaseAI is depicted in Fig.1. It comprises of three deep learning modules responsible for lung segmentation, lungs opacity detection, and chest X-ray pathology detection. The input X-ray image is resized to 256 x 256, 1024 x 1024 and 224 x 224 for lung segmentation, lung opacity detection and chest X-ray pathology classification module respectively. Lung fields identified as an output of the lung segmentation module are partitioned into six different regions and the bounding boxes that are obtained from the opacity detection model are projected on to the original chest X-ray image. The pathology detection module provides the probabilities associated with pathology labels mentioned in CheXpert dataset [14]. Finally, the outputs of all the deep learning modules are fed into the expert system to assign the COVID likelihood.

**Figure 1:**
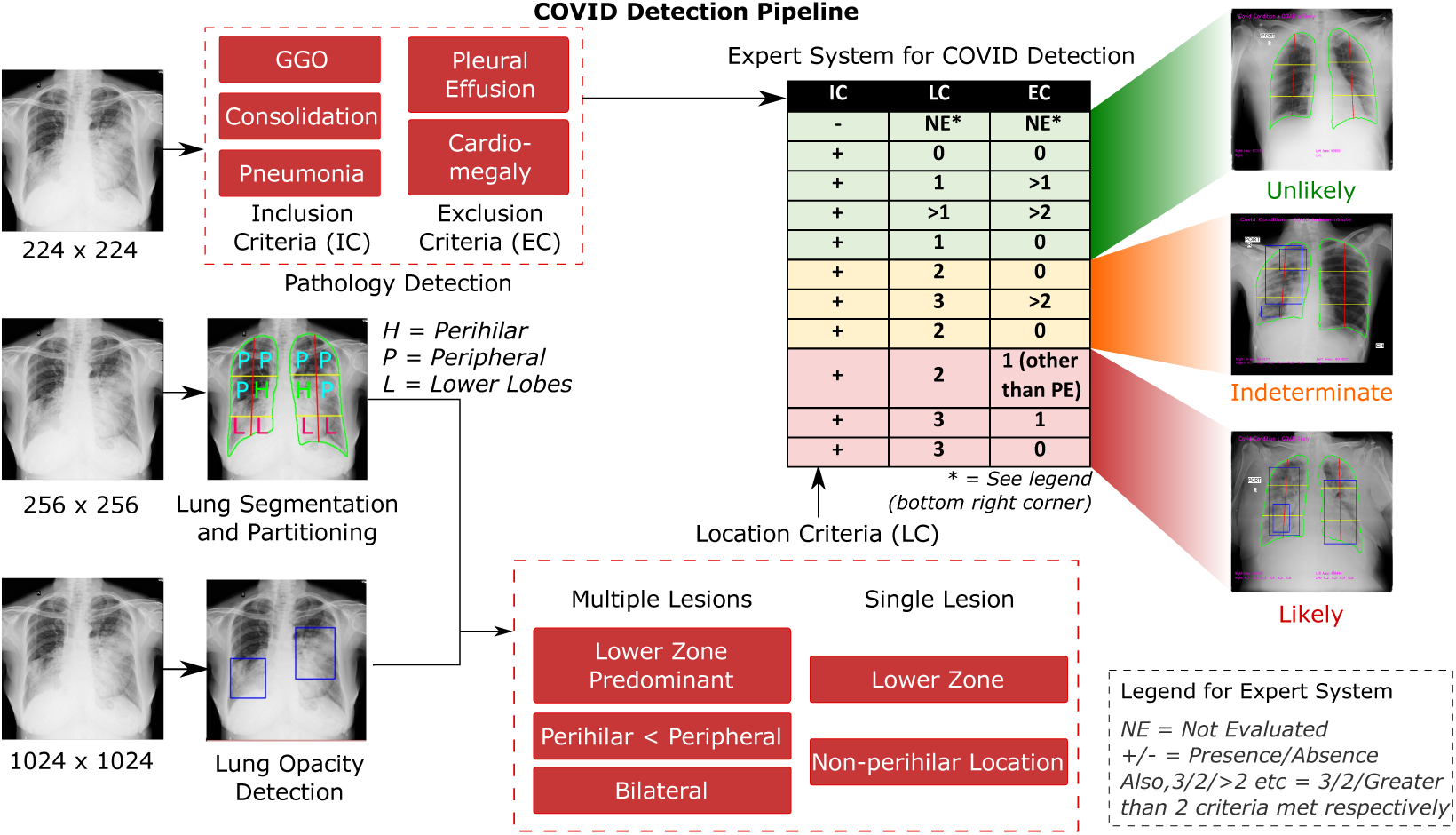
CovBaseAI model architecture for COVID likelihood Detection

### 3.1 Lung Segmentation

Lung segmentation module is a fully connected network adapted as a modification of U-Net [32]; a proven convolutional network for biomedical image segmentation. The CxR image is resized to 256 x 256 and provided as input to the lung segmentation module in order to obtain the output lung mask. U-Net being a convolutional neural network, is open to layer wise modifications and blocks of various other neural networks to enhance the segmentation output. In the U-Net architecture, we replaced the encoder part by the VGG16 [34] network which is pretrained on the ImageNet [7] dataset. For training the segmentation module, a batch size of 4 was used, learning rate was kept at .001, loss function used was BCEWithLogitsLoss, Adam was used as optimizer and it was trained for 30 epochs.

### 3.2 Lung opacity detection

Lung opacity detection is performed using a Faster R-CNN [31] architecture utilizing VGG16 network as the backbone for feature extraction in Faster R-CNN. It is a two-stage object detection network which offers high accuracy as compared to a single-stage network. It is a merger of the region proposal network (RPN) and the Fast R-CNN [9] method. In the first stage; object-like proposals are generated, second stage focuses on recognition of these proposals. Input to the Lung opacity module is the CxR image which is resized to 1024 x 1024 and the bounding boxes surrounding opacity regions are obtained as output. The hyper-parameters for opacity detection module were: for the two components in RPN, softmax classifier was used for determining the loss in score generation of each region predicted by the classification network. Smooth L1 loss was deployed to compute the loss in the regression layer, batch size was kept as 8, Adam was used as optimizer, learning rate was set to .001 and it was trained for 100 epochs.

### 3.3 Pathology detection on chest X-rays

Pathology detection on chest X-rays is performed using DenseNet-201 [13] as a backbone network. The input CxR image is resized to 224 x 224 using the linear interpolation [25] technique and was initialized with pretrained weights of imagenet dataset using transfer learning approach similar to the technique used in [26]. The pathology detection module outputs the probability corresponding to each pathology which is taken into consideration by the rule based COVID-19 likelihood detector to give final prediction. In the CheXpert dataset three labels are given corresponding to each image for each pathology i.e positive, negative, and uncertain. While training the pathology detection module; uncertainty labels were neglected and only frontal CxRs were taken into account. Hyper-parameters for training the pathology detection module were: a batch size of 32 was used, learning rate was kept at .001, sigmoid classifier was employed and it was trained for 20 epochs.

### 3.4 Lung opacity detection with lung partition

Lung mask coordinates obtained from the lung segmentation module aids us in identifying the left lung and right lung on the input CxR which are then partitioned into different regions to localize the perihilar, peripheral, middle, upper, and lower zone segments of the two lungs as this information alongwith the bounding boxes coordinates obtained from opacity detection stage would assist the rule based expert system for COVID-19 classification to predict COVID-19 likelihood.

### 3.5 Rule-based Expert Decision System for COVID classification

The rules for classification were framed by a consensus among four radiologists with more than ten years of experience in thoracic radiology. The frequency of findings on chest imaging studies including Chest X-rays and CT scans reported by several researchers were considered in establishing the consensus rules [35], [40], [12].

The rules were framed based on three broad sets of criteria: Lesion inclusion criteria, Lesion exclusion criteria, and Location criteria for lesions and pathology identified. Fig. 2 delineates the rule based inference logic utilized for defining COVID-19 likelihood. In the available literature, the algorithms have been trained to identify subtle features by neural networks which could help differentiate COVID images from Non-COVID images. We have here demonstrated an amalgamation of learning networks with experience of radiologists to have expert systems which help define the covid likelihood.

**Figure 2:**
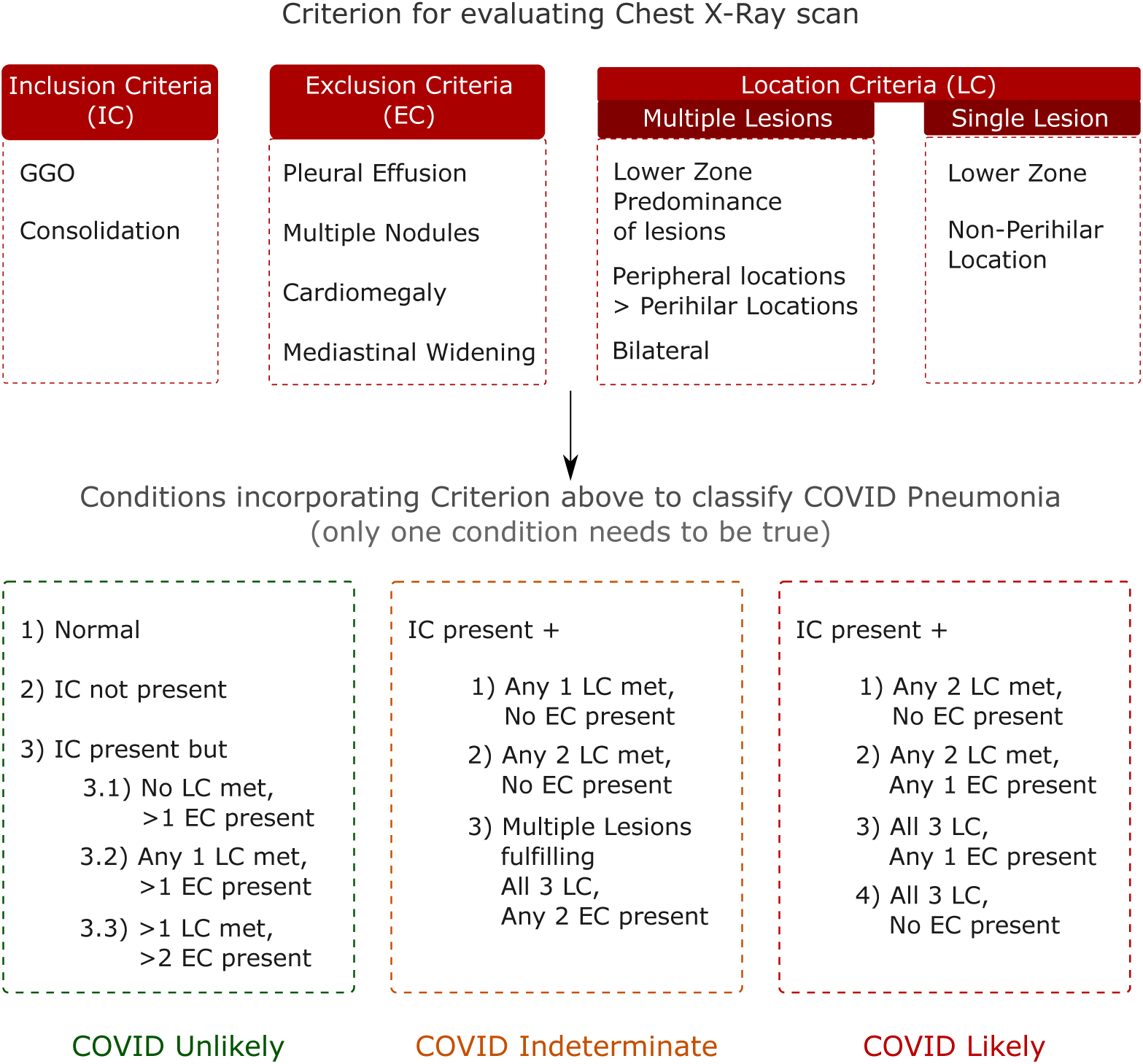
Rule based EDS for COVID inferencing devised by radiologists.

### 3.6 Validation

In order to evaluate the performance of the CovBaseAI model we used two datasets. First, 1401 randomly selected CxR from an Indian quarantine-center to assess effectiveness in excluding radiologic Cov-Pneum that may require higher care. Second, a curated data set with 434 RT-PCR positive cases of varying levels of severity and 471 historical scans containing normal studies and non-COVID pathologies, to assess performance in advanced medical settings. We compared the outputs of the model against the ground truth. Since radiological and RT-PCR ground truth vary and each has a significant error associated with it, this was done in multiple different ways as described in the dataset section and results. Generally, we prioritized radiological ground truth since that is the most relevant use case scenario at the current state of technology.

## 4 Datasets

### 4.1 Training and Validation datasets

Lung segmentation and Lung Opacity detection discussed in methodology section was performed using RSNA pneumonia detection challenge dataset [1] (for lung segmentation a subset of RSNA Pneumonia Detection Challenge dataset is used which is similar to https://github.com/limingwu8/Lung-Segmentation) in order to identify and localize pneumonia. This dataset consists of 26684 images of three different classes; Normal, Lung Opacity, and No Lung Opacity/Not Normal. Lung Opacity class consists of 6012 images. The Normal Class consists of 8851 images and No Lung Opacity/Not Normal class consists of 11821 images in DICOM format. For Lung Opacity Class, coordinates in the form of X-min, Y-min, height and width for lung opacity are provided. The size of each image is 1024 x 1024 pixels and Lung Opacity class contains 9555 coordinates of lung opacities in 6012 images. Pathology detection on chest X-rays was carried out using CheXpert [14] dataset which was released by Stanford ML group in 2019 as competition for automated chest X-ray interpretation. CheXpert dataset consists of following pathologies: Lung Lesion, Edema, Consolidation, No Findings, Atelectasis, Pneumothorax, Pleural Effusion, Pneumonia, Pleural Others, Cardiomegaly, Enlarged Cardiomediastinum, Lung Opacity, Fracture and Support Devices. For training and validation of pathology detection module only frontal CxRs were used and CxRs with uncertain labels were not used.

### 4.2 Independent validation datasets

To determine usefulness as a screening and triage tool, we used 1401 CxRs (IITAC1.4K) obtained from an Indian quarantine centre. SARS-CoV-2 infection status was not known for this dataset, but this reflects an actual mix of cases likely to present to quarantine centers where a determination has to be made regarding patients that can be safely kept in general isolation and those that may need medical care. This dataset is part of over 8500 CxRs from NSCI Dome initiative at Mumbai. Of these 1401 CxRs were selected randomly and annotated carefully by an experienced radiologist for the presence of various pathologies. This dataset is composed of 135 and 1266 COVID-19 (+ve and -ve respectively) CxRs. (available at http://covbase4all.igib.res.in)

To determine usefulness as a diagnostic tool for SARS-CoV-2 infection or for confirmed COVID-pneumonia, we used a validation dataset comprised of a total of 905 CxRs with 434 RT-PCR positive scans and 471 historical scans that were acquired before the worldwide outbreak of COVID-19. The X-rays were annotated with the consensus of four senior radiologists on CARPL platform provided by Centre for Advanced Research in Imaging, Neuroscience, and Genomics (CARING), India for the presence or absence of consolidation at the study level. A snippet of CARPL platform used for annotation and validation is depicted in Fig.3. The presence or absence of Pleural Effusion, Atelectasis, Cardiomegaly, Fibrosis, Mediastinal Widening, Nodule, Pleural Effusion, and Pneumothorax were also recorded. The CovBaseAI model was validated on this set of 905 CxRs using the following levels of tests:

**Figure 3:**
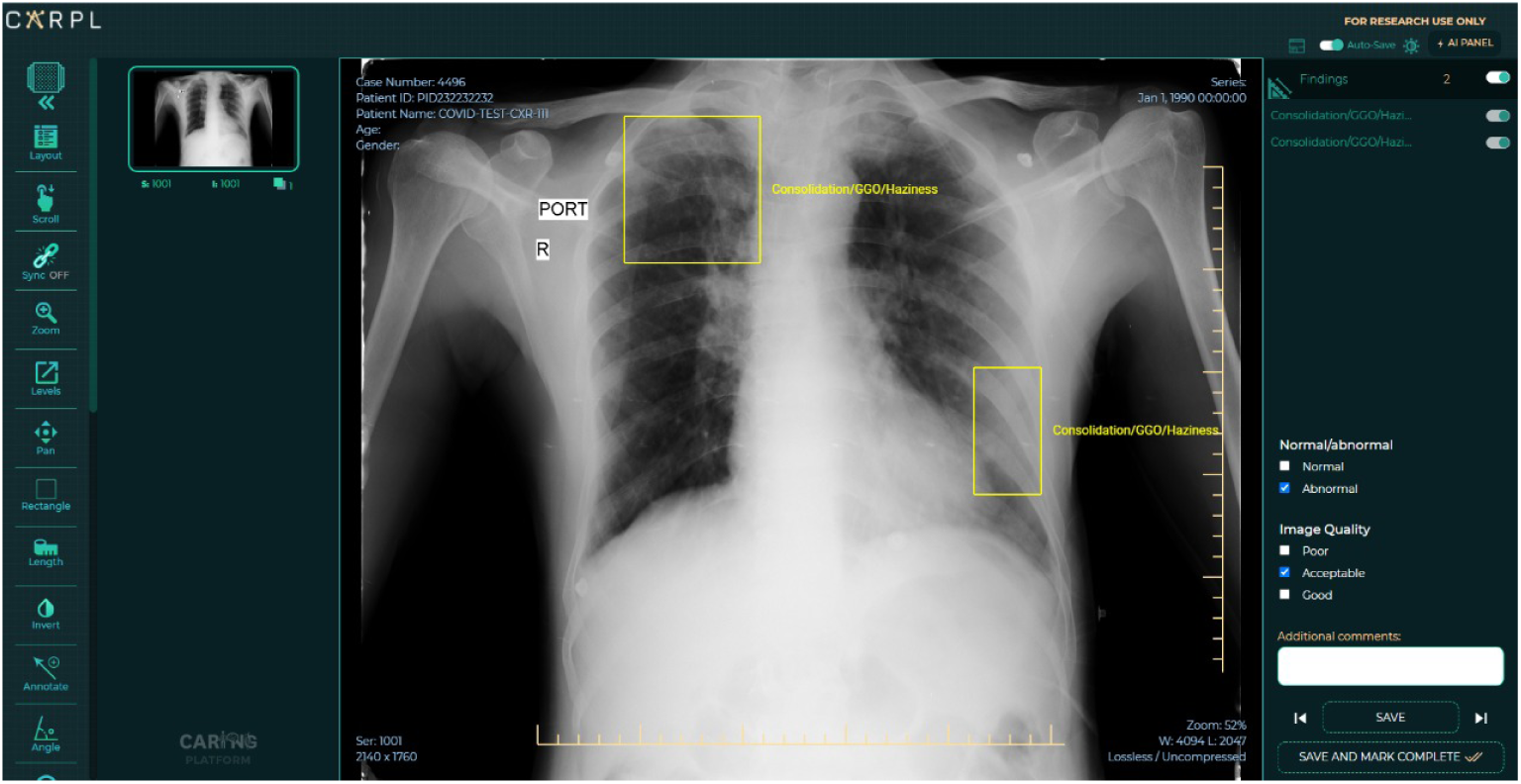
A snippet of CARPL platform used for annotation and validation. *(Image courtsey: CARING, India)*

1. **Pneumonia Detector (PD1K):** Prediction output of the CovBaseAI model was compared against the pneumonia/consolidation label annotated by radiologists (irrespective of RT-PCR status). PD1K consists of 484 +ve and 421 –ve labels.
2. **COVID Infection Detector (CID1K):** COVID status predicted by CovBaseAI model was compared against the RT-PCR results for testing its ability to detect COVID infection. CID1K consists of 434 +ve and 471 -ve labels.
3. **COVID Pneumonia Detector (CPD600):** All cases with RT-PCR positive and radiologist annotated consolidation positive cases were taken as a positive class [336 cases] and all RT-PCR negative & consolidation negative cases [323 cases] were taken as negative class.

Additionally, the CovBaseAI model was also validated on a test dataset as mentioned in [36]. The instructions for creating the dataset can be found at (https://github.com/ddlab-igib/COVID-Net/blob/master/docs/COVIDx.md). The aforementioned test dataset is referred as COVIDx1K in the manuscript. At the time of writing this manuscript, COVIDx1K comprises of 885 normal and 100 COVID-19 CxRs (test images with pneumonia label were not considered in COVIDx1K dataset).

## 5 Results

The segmentation algorithm of lung mask detection was validated on 100 chest X-rays as test set, from the pool of 1000 X-rays from RSNA dataset. We obtained 0.91 as Jaccard similarity index [8] on the validation set. Accuracy/Loss plot on training and validation data has been depicted in Fig.4a.

The lung opacity detection module was validated on 1012 test X-ray images from RSNA Kaggle dataset. The exactness of object detection is usually well determined by mAP (Mean Average Precision) [19], for opacity detection mAP of 0.34 is achieved. Based upon the pathology detection module’s classification probability scores corresponding to pathologies included in CheXpert dataset, the AUC’s (Area under the ROC Curve) for Cardiomegaly, Consolidation, Lung Opacity, No Findings, Pleural Effusion and Pneumonia was 0.83, 0.90, 0.90, 0.91, 0.93, 0.78 respectively. The ROC curves for pathology detection module for different pathologies on the CheXpert validation set has been depicted in Fig.4b.

**Figure 4:**
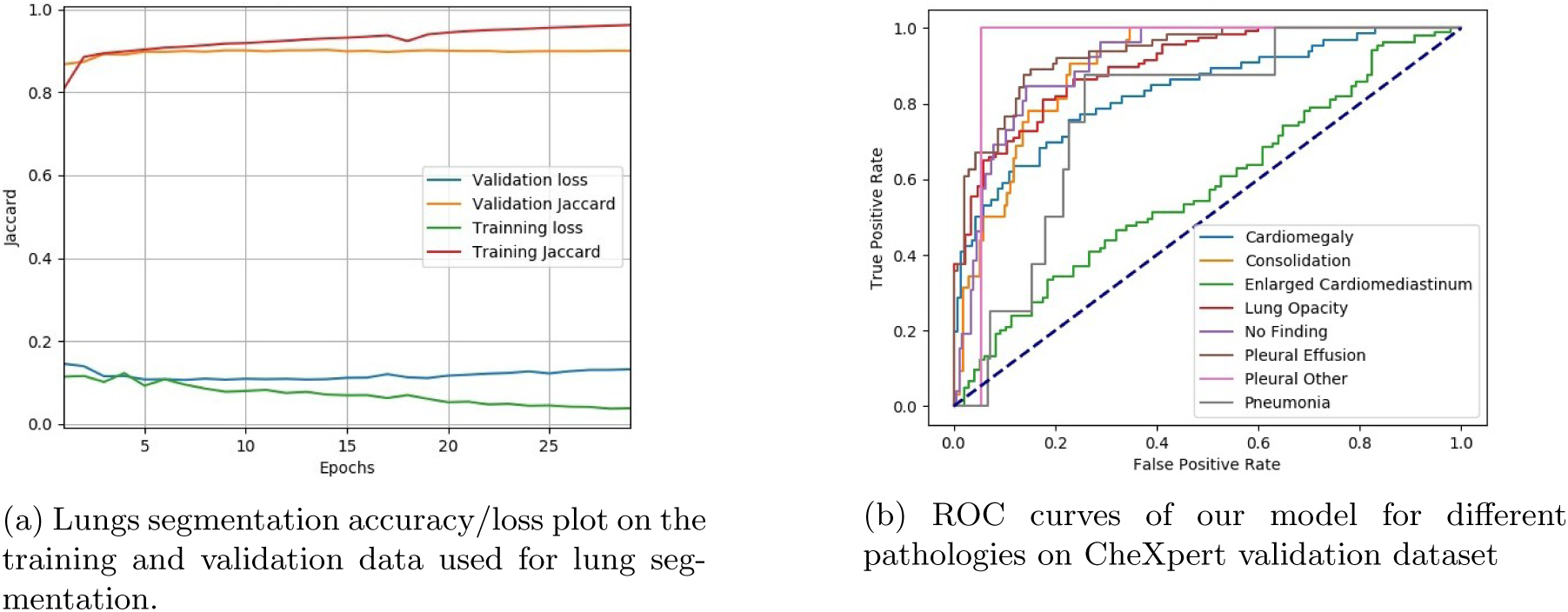
Results of Lung segmentation and Pathology detection module

Table.1 depicts the performance metrics of CovBaseAI model on different independent validation datasets. Since the ground truth available for validation dataset was either positive or negative(i.e. RT-PCR +ve/-ve or consolidation +ve/-ve) and output of CovBaseAI model is given in three classes (COVID likely, COVID indeterminate and COVID unlikely), the COVID indeterminate class was merged with COVID likely class for calculating performance metrics such as Sensitivity, Specificity, Accuracy, Negative Predictive Value (NPV) etc. Fig.5 depicts the sample of True positive, False positive, True negative and False negative obtained by CovBaseAI model. Validation studies corresponding to the CovBaseAI model was done using CARPL. Fig.6 shows bounding boxes of representative false positive images from IITAC1.4K data that were read as normal by the radiologist. On review, the findings inside these bounding boxes were prominent bronchovascular markings, a common finding in the Indian subcontinent, with no clinical significance. Table.2 shows the concordance of bounding boxes (mAP) between lesions identified by AI and radiologists. In 905 CxRs (434 RT-PCR +ve and 471 historical scans) from Independent Validation datasets, read by four radiologists, intersections between AI-human pairs are similar to human-human pairs. Further, in case of IITAC1.4k dataset which is read by a single radiologist, majority of the time the bounding box of AI and radiologist intersect. Thus the determination of COVID-19 pneumonia in our model is based on the same parts of the CxR as human radiologist and explainability can be considered to be high.

To understand the relative performance of our model versus a publicly available COVID-19 CxR algorithm, COVID-CAPS that has reported high performance characteristics on a subset of data used in [36], we deployed COVID-CAPS on the Indian dataset. The sensitivity and specificity were only 33% and 67% on IITAC1.4K data that represents an actual use case scenario. The accuracy was below chance (50%) on the CPD600 dataset, which is more challenging. This illustrates the difficulty in portability of DL solutions to other regions. In contrast, CovBaseAI performed well across datasets, with higher specificity on a western dataset (COVIDx1K) than Indian ones, which reflects the fact that the DL components had never seen Indian data.

**Figure 5:**
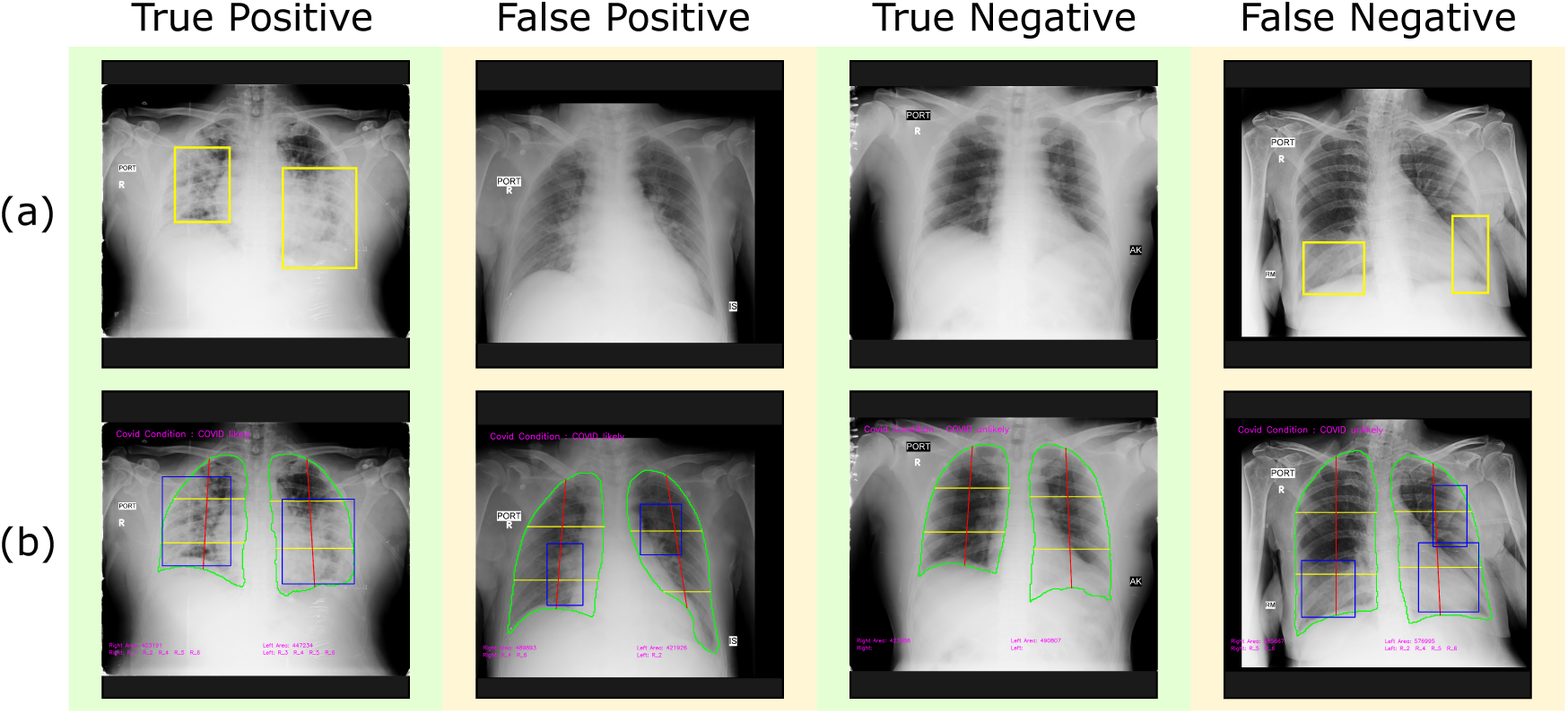
Samples of true positive, false positive, true negative and false negative from independent validation set of 905 CxRs (a) Ground truth CxR (b) CovBaseAI inferencing result.

## 6 Discussion

There is a great need for AI solutions that have explainability and an assurance of a minimum performance when exposed to data they have not been trained on. While COVID-19 has brought out this need globally, not discriminating between the developed and developing world, the fact of the matter is that this problem has been around for a long time. Here, we have addressed the problem of diagnosing COVID-19 pneumonia in Indian CxR without training on either COVID-19 or Indian CxR data, by using an explainable system driven by logic and expert opinion. While the solution has useful performance characteristics (see Table 1) across Indian (IITAC1.4K, PD1K, CPD600) and western (COVIDx1K) datasets, with high explainability (see Table 2), the insights lie in the differences. In particular, 97% specificity on a western dataset, but 86% specificity on a comparable Indian dataset is notable.

**Table 1:**
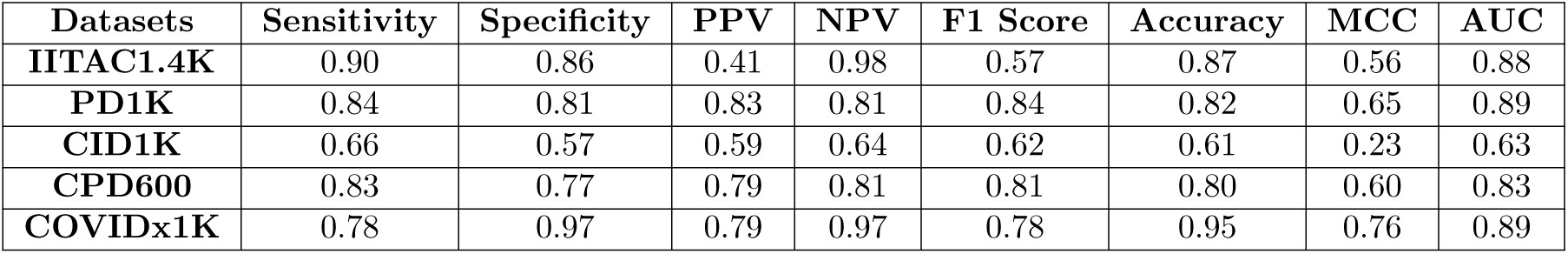
Performance metrics of CovBaseAI model on independent validation datasets. PPV=Positive Predictive Value; NPV=Negative Predictive Value; MCC=Matthews Correlation Coefficient; PD1K=Pneumonia Detector; CID1K=Covid Infection Detector; CPD600=Covid Pneumonia detector.

| Datasets | Sensitivity | Specificity | PPV | NPV | F1 Score | Accuracy | MCC | AUC |
| --- | --- | --- | --- | --- | --- | --- | --- | --- |
| IITAC1.4K | 0.90 | 0.86 | 0.41 | 0.98 | 0.57 | 0.87 | 0.56 | 0.88 |
| PD1K | 0.84 | 0.81 | 0.83 | 0.81 | 0.84 | 0.82 | 0.65 | 0.89 |
| CID1K | 0.66 | 0.57 | 0.59 | 0.64 | 0.62 | 0.61 | 0.23 | 0.63 |
| CPD600 | 0.83 | 0.77 | 0.79 | 0.81 | 0.81 | 0.80 | 0.60 | 0.83 |
| COVIDx1K | 0.78 | 0.97 | 0.79 | 0.97 | 0.78 | 0.95 | 0.76 | 0.89 |

**Table 2:** Bounding box analysis on 905 CxRs(434 RT-PCR +ve and 471 historical scans) from Independent Validation datasets. RAD=Radiologist; mAP=Mean Average Precision

|  |  | Prediction Box |  |  |  |  |
| --- | --- | --- | --- | --- | --- | --- |
|  |  | CovBaseAI (mAP) | RAD 1 (mAP) | RAD 2 (mAP) | RAD 3 (mAP) | RAD 4 (mAP) |
| Ground Truth | RAD 1 | 0.78 | 1 | 0.79 | 0.54 | 0.79 |
|  | RAD 2 | 0.73 | 0.80 | 1 | 0.51 | 0.75 |
|  | RAD 3 | 0.43 | 0.54 | 0.50 | 1 | 0.46 |
|  | RAD 4 | 0.81 | 0.81 | 0.75 | 0.47 | 1 |

Most AI solutions, large datasets for training, quality annotations, have come from the developed world. Yet, it is well known that even something as simple as CxR look different in regions with high ambient pollution, exposure to dirty fuel, endemic tuberculosis, low body mass, and other differences in lifestyle [4]. It is well known in the radiology community that this well described “dirty lungs” appearance with prominent bronchovascular markings can easily be confused with abnormal. AI systems, however, are likely to give false positives when exposed to such images, if not previously exposed during training (see Figure 6 for examples). Such false positives have been part of abstracts and anecdotal experiences [16] but publication bias against failures has led to a situation where AI solutions for interpreting CxR are working very well in published manuscripts, without similar enthusiasm in actual practice. The only long-term solution is to have high quality annotated images from LMIC regions, via global initiatives. In the short-term, use of human logic filters can help in keeping the specificity high enough for clinical use. In our results, the specificity was 97% on a western dataset and 86% for India quarantine center data. A pure DL algorithm that reportedly performed well on a subset of the western data, had only 67% specificity on the quarantine center dataset, which makes it unsuited for clinical use in India. Notably the human logic filters did not substantially compromise sensitivity, which was in the 80-90% zone for COVID-pneumonia, falling to 66% for COVID infection. While it does not seem possible, using our approach, to detect SARS CoV2 infection, without pneumonia, this likely reflects a fundamental lack of sensitivity of CxR rather than a flaw in the algorithm, and is consistent with WHO guidelines that do not recommend imaging for infection status determination [39]. The performance characteristics across all the COVID pneumonia datasets are in the clinically useful range and AUC was well above 0.8, the usual threshold at which diagnostic tools are considered useful. Further, the explainability was high (Table 2) and led to an understanding of the underlying issues (Figure 6), which makes further improvements possible. Such work is now underway and will hopefully lead to more usable AI systems for the Indian subcontinent and similar global regions.

**Figure 6:**
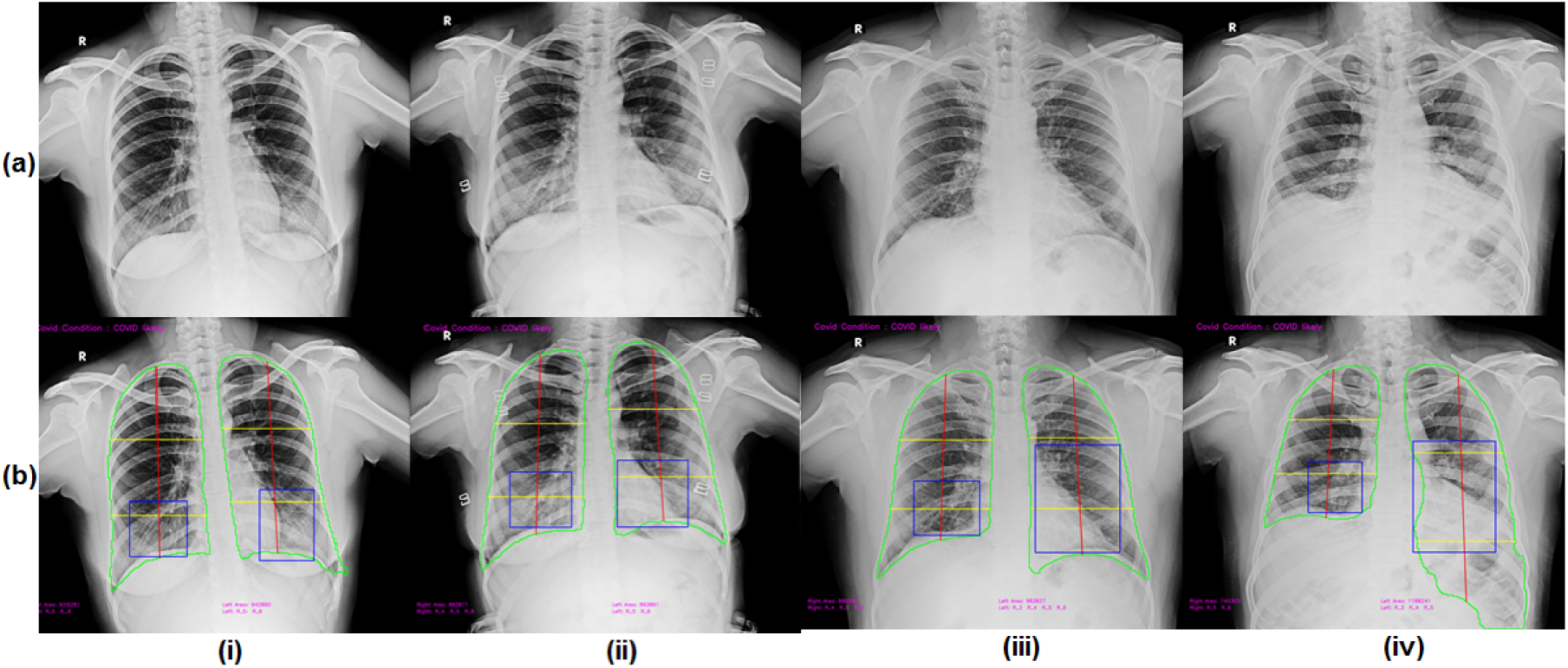
Samples of false positive from IITAC1.4K dataset (a) Ground truth CxR (b) CovBaseAI inferencing result.

## 7 Conclusion

Several Deep learning architectures have been proposed for COVID-19 detection from CxRs. Most of these methods used limited COVID-19 datasets for training purposes. Models trained on these limited datasets are not ready for real world usage without training on a large variety of regional datasets to accommodate local variations of data acquisition hardware, population specific differences and temporal disease manifestation. For physicians to be able to use AI solutions for screening or triaging, the tools should provide robust detection results, across global data, with explainable findings and grading of severity, but requiring minimal amounts of new data so that solutions can be rapidly developed and deployed. Such an all inclusive tool is yet to be developed for COVID-19, but CovBaseAI is a useful step in that direction.

## Data Availability

The datasets used are available at https://covbase4all.igib.res.in/

## Data Availability

The datasets used are available at https://covbase4all.igib.res.in/

## 8 Author Contribution

PSG, SSP, APS, SP, AS, SS, SSingh and ASM contributed for the model development. PS, NB, VS, VM, VV and DD contributed for model evaluation and validation. RMV, RT and SG contributed for model building platform support. VV, AnjaliA, AK and HM contributed as radiologists for the ground truth creation and rule based logic for EDS. DD, VM and AnuragA conceptualized the idea and coordinated the research activity. PSG, SSP, APS, ASingh, PS, NB, VS, VV, AnuragA, AnjaliA and DD contributed in writing and editing of the manuscript. All authors reviewed and agree with final version of the manuscript.

## 9 Funding

This activity is partially funded by project MLP-2002 (CSIR-IGIB) and CSIR-Intelligent Systems Mission Mode Project (CSIR-CEERI) of Council of Scientific and Industrial Research (CSIR), India.

## 10 Competing Interests

CARPL is a commercial product owned by CARING (Mahajan Imaging) and was utilized for this work as a part of R&D MoU between CSIR-IGIB and Mahajan Imaging.

## 11 Acknowledgments

All the authors duly acknowledge IIT Alumni Council for their contribution of 8500 CxR data and Drs. Swarn Kanta and Virender Kumar Jain for radiologist and physician inputs on the 1401 CxRs as a subset of the above data.

## Notes

### Author Declarations

Institute Human Ethics Committee, CSIR-IGIB, New Delhi Ref# - CSIR-IGIB/IHEC/2020-21/02 Dated - 28/03/2020

